# Vaccine Stockpile Sharing For Selfish Objectives

**DOI:** 10.1101/2022.04.28.22274446

**Authors:** Shashwat Shivam, Joshua S. Weitz, Yorai Wardi

## Abstract

The COVAX program aims to provide global equitable access to life-saving vaccines. However, vaccine protectionism by wealthy nations has limited progress towards vaccine sharing goals. For example, as of April 2022 only *∼*20% of the population in Africa has received at least one COVID-19 vaccine dose. Here we use a two-nation coupled epidemic model to evaluate optimal vaccine-sharing policies given a selfish objective: in which countries with vaccine stockpiles aim to minimize fatalities in their own populations. Despite the selfish objective, we find it is often optimal for a donor nation to share a significant fraction of its vaccine stockpile. Mechanistically, sharing a vaccine stockpile reduces the intensity of outbreaks in the recipient nation, in turn reducing travel-associated incidence in the donor nation. This effect is intensified as vaccination rates decrease and epidemic coupling increases. Despite acting selfishly, vaccine sharing by a donor nation significantly reduces transmission and fatalities in the recipient nation. Moreover, we find that there are hybrid sharing policies that have a negligible effect on fatalities in the donor nation compared to the optimal policy while significantly reducing fatalities in the recipient nation. Altogether, these findings provide a rationale for nations with extensive vaccine stockpiles to share with other nations.

## Introduction

Vaccines have been shown to be effective in reducing severe infections, hospitalizations and fatalities [1, 2], however, the availability of doses across nations remains highly non-uniform [3]. While sharing vaccines between nations is now seen as a global imperative [4, 5, 6], most wealthy nations have not initiated vaccine sharing campaigns that would promote vaccine equity in neighboring countries and/or worldwide. Vaccine sharing can reduce the intensity of outbreaks and reduce the risk of case importation when stringent regulations are relaxed in a donor country [7, 8, 9]. However, vaccine sharing comes with a cost: reducing the availability of vaccines for the donor nation. This reduction is perceived to lead to greater infections, hospitalizations, and fatalities in donor countries – and may also come with political costs for decision-makers who propose seemingly altruistic policies to share their vaccine stockpiles.

Epidemic models can be used to assess the costs and benefits of vaccine stockpile sharing between nations. A recent paper [10] studied the efficacy of vaccine sharing between two nations with extensive travel between the donor and recipient nation. Epidemic dynamics within each country were coupled through the occasional importation of cases and/or the travel of individuals to and from the donor country to the recipient country (where they could be infected) and back again (where they could accelerate disease transmission). This study concluded that the greatest reduction in fatalities within the vaccine-rich (donor) nation occurred in the absence of sharing. This epidemic model analysis included a time-dependent sharing protocol while assuming that vaccination rates were high, e.g., the eligible population would be fully vaccinated within 50 days. Related efforts evaluated the question of optimal vaccine sharing after a donor nation is already at or near herd immunity [11]. In that case, donating vaccines to the group of COVAX countries can reduce fatalities within the donor nation. In doing so, this study views donation after herd immunity as a binary alternative, either donating all the ‘surplus’ vaccines to a recipient (vaccine-poor) country or using the vaccines internally. In both cases, the implicit assumption is that vaccination rates in donor countries are fast and can rapidly reach herd immunity. However, in practice, vaccination coverage has increased far slower than anticipated, e.g., in the US, only 60% of the population has been fully vaccinated more than one year after the widespread availability of vaccines [12]. This failure to rapidly vaccinate populations in vaccine-rich countries raises new questions on how to optimally utilize vaccine stockpiles to reach selfish objectives.

In this manuscript, we consider a two-nation epidemic model in which a donor (vaccine-rich) country can share vaccines with a recipient (vaccine-poor) country. In doing so, we seek the optimal sharing policy given a selfish objective - reducing fatalities only within the donor country. Further, we assume that the population is immunologically naive - given our intent to establish the prudence of prior decisions as well as the potential for improved decision-making in the future. In the present analysis, we consider a broader range of vaccination rates than has been considered in related studies. This choice is motivated by realized coverage rates from the USA (where ~62% of the population was fully vaccinated in a period of 1 year) and UK (~69% were fully vaccinated in 1 year) [13]. Despite the selfish objective, we find that there is a broad range of vaccine uptake rates and cross-nation epidemic coupling rates in which the optimal policy is for the donor nation to share a substantial fraction of its vaccine stockpile with a recipient nation. As we show, sharing vaccines can reduce the fatalities in the donor country (acting in its own self interest) with the added benefit that acting selfishly induces sharing that also curbs the spread of COVID-19 in the recipient nation.

## Vaccine sharing problem

We consider two countries, A and B, each confronting a COVID-19 outbreak in which one country (A) has a vaccine stockpile and the other country (B) does not. The outbreak is modelled using SEIRV (Susceptible - Exposed - Infected - Recovered - Vaccinated) dynamics. The system dynamics of the countries are coupled and active infections in one country can cause infections in the other country. Further, country A has the option of donating a part of its vaccine stock to country B. This vaccine sharing can only be done once at the start of the outbreak - between A (the donor country) and B (the recipient country).

The objective of the selfish, optimal vaccine sharing policy is to minimize fatalities in the donor country. The epidemic dynamics between countries are coupled. Here, we explore optimal policies as a function of epidemiological parameters as well as two features of the two-nation problem: (i) the value of the epidemic-coupling constant; (ii) the rate of vaccine uptake in the donor country. An increase in the epidemic-coupling constant makes it more likely that infections in the recipient country lead to new cases in the donor country (and vice-versa). The vaccine uptake rate controls the rate at which a donor country can potentially use its vaccine stockpile; and we focus on limits in which the rate of vaccine uptake (on the order of a year) is slower than that of typical outbreak dynamics (i.e., on the order of months). In all cases the objective of the donor nation is to minimize fatalities in its own country. Later we relax this assumption to propose near-optimal solutions that result in substantially-lower mortality rates in the recipient country with only small increases in the morbidity experienced by the donor nation. The details of the dynamic model and related optimization problem are available in the Methods section.

## Results

We consider the two countries to have an initial population of 10^7^ with 500 initial infections in which country A has enough vaccines to fully vaccinate 7 × 10^6^ individuals. The optimal vaccine sharing fraction *µ*\* is the value which minimizes the total fatalities in A over a given time horizon, regardless of its effect on the morbidity or death rate in B. We explore the dependence of the optimal policy with respect to the daily vaccination rate, *λ*, and the epidemic coupling constant, *κ*. Simulation results are contained in the heatmap shown in Fig. 1. For *κ* ≤ 10^−6^, the optimal vaccine sharing fraction is 0 if the vaccination rate is sufficiently high. These findings are consistent with the results reported in [10] - the rationale is that it is more effective to rapidly immunize nearly the entire population in county *A* than to share and reduce the importation of cases. However, when vaccination rates within A are low (e.g., less than 0.19%*/*day, equivalent to 70%*/*year), then the optimal vaccine sharing fraction is positive for all epidemic coupling constants studied. When the epidemic coupling constant between the two countries is stronger, then it is beneficial to donate more vaccines to country B. From the heatmap, we find that the optimal fraction is negatively correlated with the vaccination rate. Moreover, we also find a critical transition: if the vaccination rate is low, then the optimal policy is to share vaccines with country B even for very low epidemic coupling constant values.

**Figure 1:**
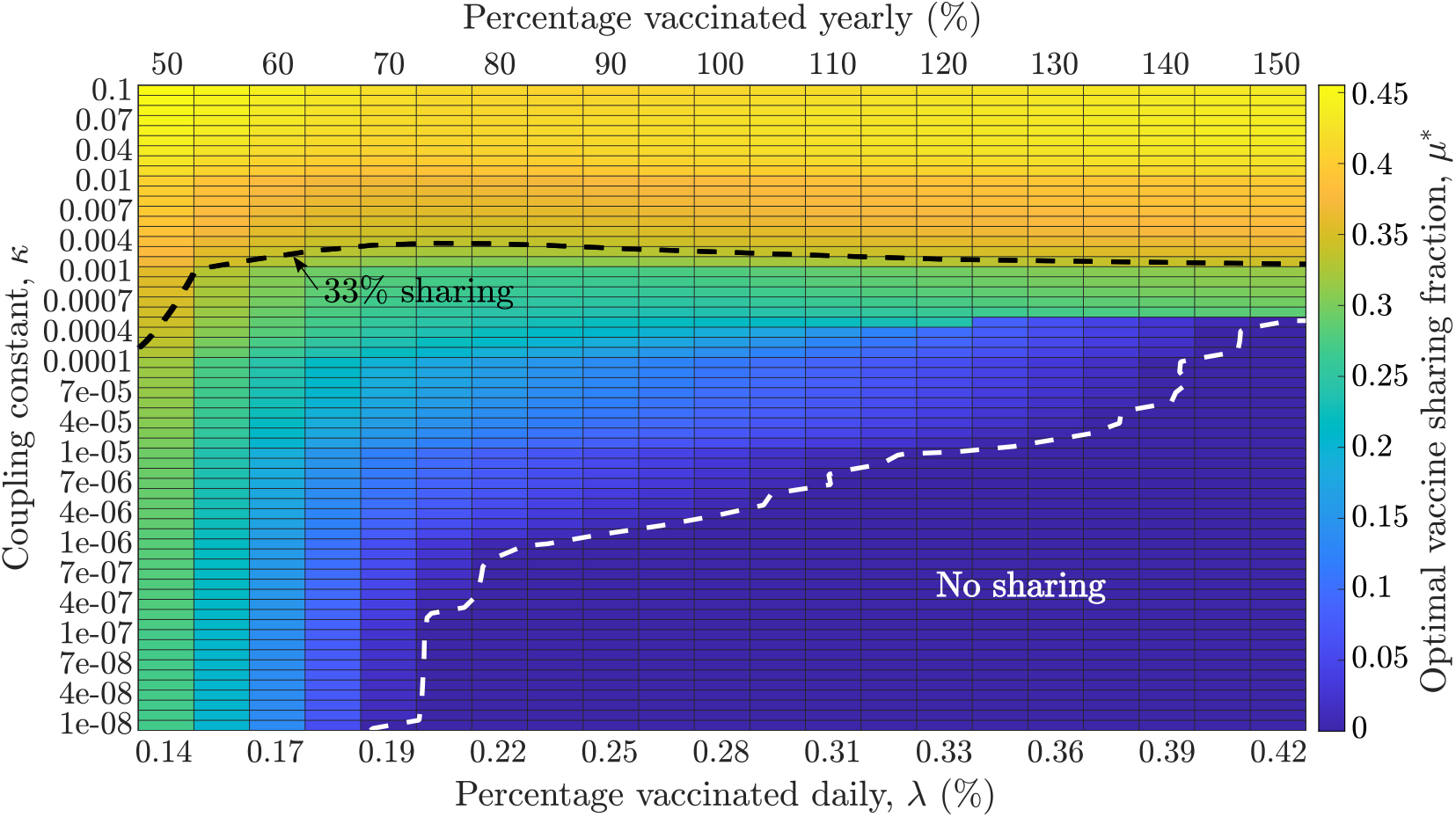
Dependence of the optimal fraction of vaccines (*µ*\*) to be donated as a function of the vaccination rate, *λ* (x-axis) and the epidemic coupling coefficient, *κ* (y-axis). For very low values of *κ* and moderate to high vaccination rates, the optimal sharing fraction is 0. For all other values, the optimal vaccination donation fraction from country A to country B is positive, increasing with the epidemic coupling constant and the realized vaccination rate in country A. The black dashed line shows a level curve of *µ*\* = 1*/*3, while the white dashed line demarcates the region where *µ*\* = 0, that is, there is no vaccine sharing.

Fig. 1 provides the optimal vaccine sharing fraction for different settings, however, it does not quantify the efficacy of implementing such a policy on epidemic outcomes. To do that we quantify the fatalities in A and B when no vaccines were shared, namely *µ* = 0, and compare these baseline levels of fatalities to simulated epidemic outcomes for different values of *µ*. Fig. 2 shows the deaths per 10^7^ individuals in both countries for different values of *µ*, for three values of the epidemic coupling constant *κ* (low, medium and high), with a fixed daily vaccination rate of 0.277% of the total population. For the three values of *κ*, sharing a small fraction of the vaccines (*µ >* 0) rapidly reduces the fatalities in B and either decreases the fatalities in A (*κ* = 10^−2^) or has a negligible adverse effect (*κ* = 10^−4^, 10^−6^). For *κ* = 10^−6^, the optimal sharing fraction is *µ*\* = 0, hence the optimal policy is the same as the no-share policy. For *κ* = 10^−4^, the fatality reduction in A is negligible, but there is a ~24% reduction in fatalities in B given an optimal sharing fraction of *µ*\*-0.11. Lastly, for *κ* = 10^−2^, the optimal policy is to share *µ*\*-0.38, or more than one-third of the donor nation’s vaccine stockpile. Compared to the no-share policy (*µ* = 0), there is a -50% and -90% reduction in fatalities in countries A and B respectively. By donating a part of the vaccine stock to B, the infections of individuals in country B are reduced which in turn reduces the cross-infections of individuals in country A. However, this level of vaccine sharing also reduces the fraction of the population of country A which is vaccinated. The magnitude of these competing factors is dependent on the epidemic coupling constant *κ*, which explains the shift towards a higher sharing policy as *κ* increases.

**Figure 2:**
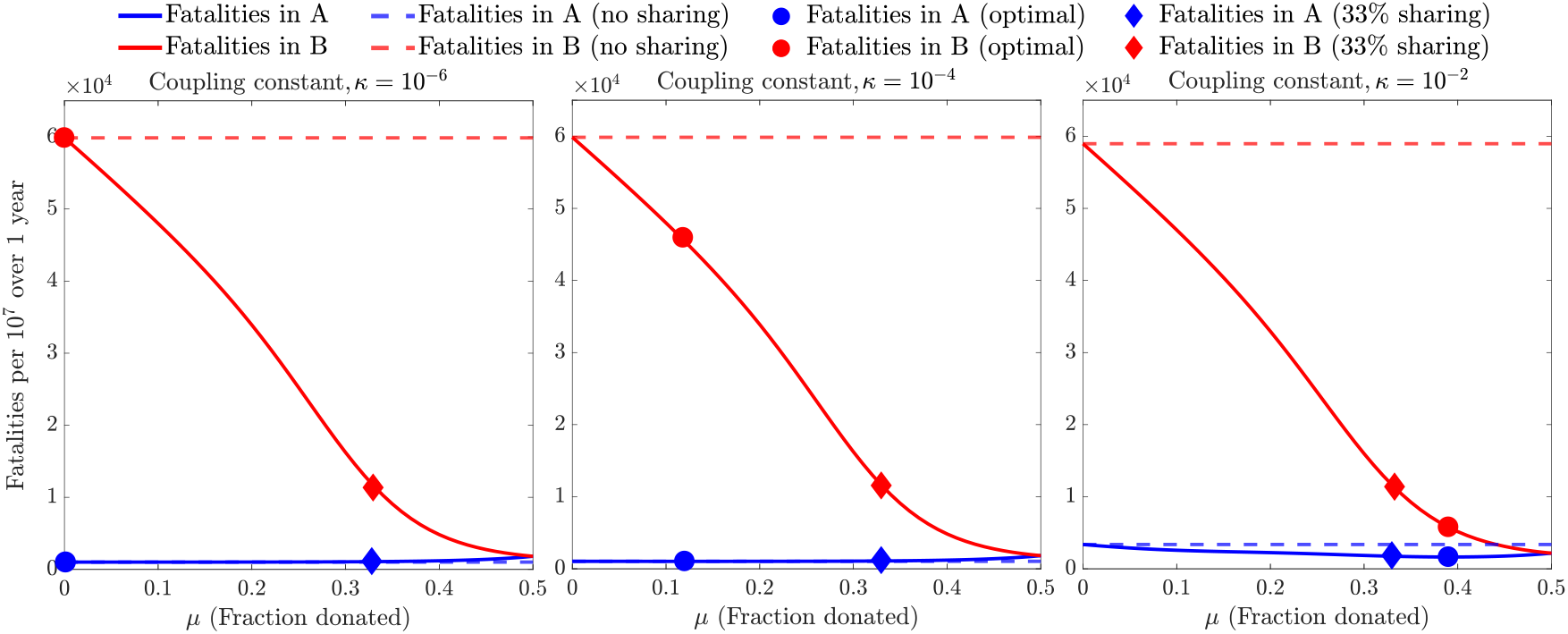
Model-based fatalities per 10^7^ over 1 year in countries A and B for different values of vaccines donated from A to B (*µ*). The value of *µ* which minimizes the fatalities in country A is termed the ‘optimal policy’. The simulation is run for low, medium and high epidemic coupling constants, *κ* (with *κ* {∈ 10^−6^, 10^−4^, 10^−2^}) and the daily vaccination rate is held fixed at 0.28% of the total population in all three panels. The optimal fraction for every *κ* is marked on the corresponding plot with a circle, along with the fatalities in countries A and B for a 33% sharing policy (diamond) and the no-share policy (dashed line).

**Figure 3:**
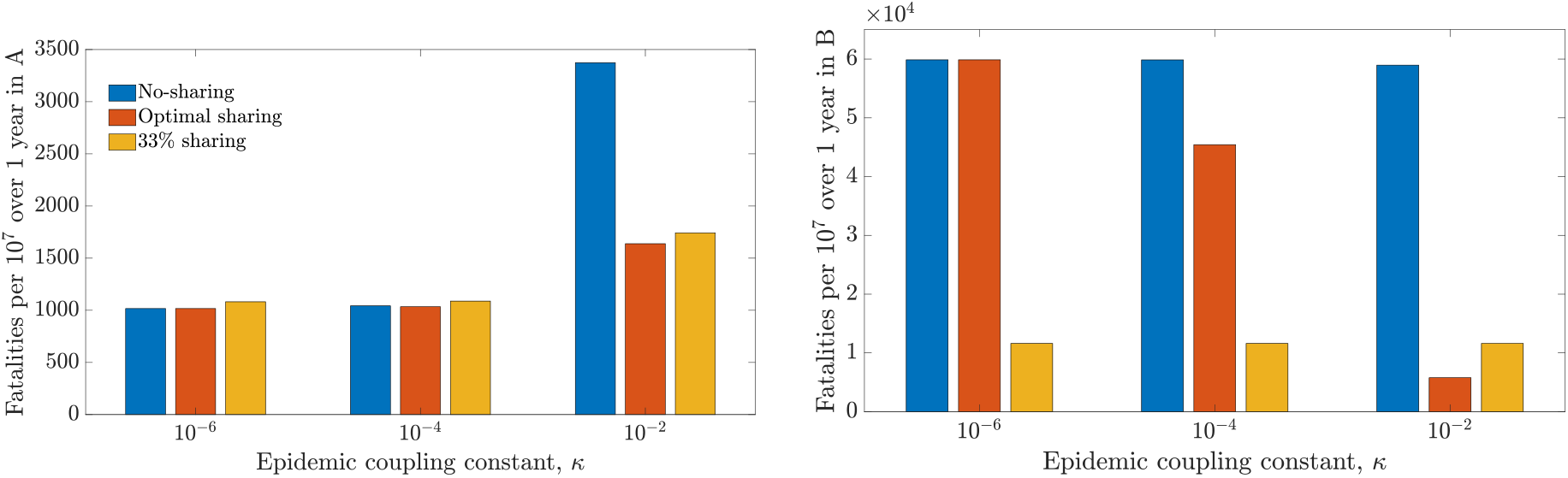
Fatality per 10^7^ over 1 year in country A and B respectively for different sharing policies - no-sharing, optimal and 33% sharing. The daily vaccination rate is set as 0.28% of the total population. The two bar graphs list the total fatalities for all three policies (no-sharing, optimal and hybrid) shown in Fig. 2.

This optimal-sharing analysis revealed that small deviations from the optimal policy led to small changes in fatalities in A in both the high and medium epidemic coupling scenarios (see Fig. 2). This suggests that using a higher-than-optimal fraction (when *µ*\* is either zero or a small fraction) would likely have negligible effects on country A, but confer significant benefits on country B. Based on this observation, we propose a hybrid policy, which is the same as the optimal policy when *µ*\* is high, and has a fixed, non-zero vaccine sharing fraction when *µ*\* is low. Denoting by 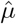 the vaccine-sharing fraction resulting from this policy, we define it as 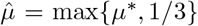. To examine the efficacy of the optimal and hybrid policies, we compare the reduction in fatalities in countries A and B using these policies against the no-sharing policy. Fig. 4 compares the hybrid, optimal and no-sharing policies, revealing that the hybrid policy increases fatalities in A by less than 10% when the vaccination rate is high (≥75%) and the epidemic coupling constant is low. However, for these scenarios, there is a large reduction in fatalities in B (≥70%), which is greater than the corresponding reduction achieved by the optimal policy. Thus, by slightly relaxing the requirement that A have a strictly selfish policy, vaccine sharing can lead to dramatic changes in fatalities in the recipient country with limited impacts on the donor country. Similar results are found on a continuum of scenarios when varying both the vaccine uptake rate and the epidemic coupling constant (see Figs. S1 and S2).

**Figure 4:**
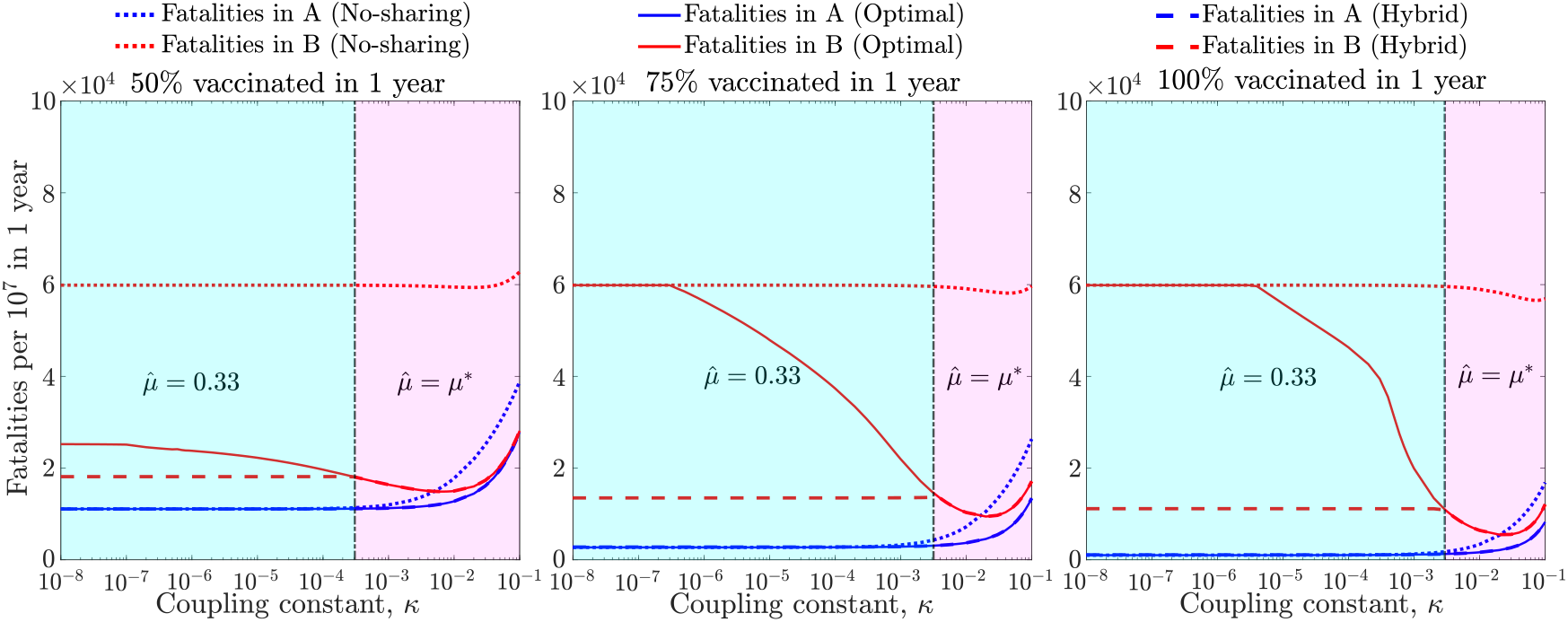
Fatalities per 10^7^ over 1 year for the no-sharing, optimal and hybrid policies in countries A and B given different epidemic coupling constants (*κ* ∈ [10^−8^, 10^−1^]) and three vaccination rates (50%, 75% and 100% of the population vaccinated in 1 year). The hybrid policy maintains a vaccine sharing fraction of 0.33 to the left of the vertical black line, where the optimal vaccine sharing fraction is lower than 0.33 (shown here in cyan). To the right, the optimal vaccine sharing fraction is equal or greater than 0.33, so the hybrid vaccine sharing fraction is equal to the optimal in this region (shown here in magenta). The interpretation of the curves is same for each of the panels: blue denotes fatalities in A and red denotes fatalities in B. For each, the results are shown for no-sharing (dotted), optimal sharing (solid) and hybrid (dashed).

## Discussion

The travel between countries experiencing an epidemic can lead to enhanced transmission and mortality. In order to evaluate the impact of vaccine sharing on epidemic outcomes, we explored the optimal sharing policy between a donor country and a recipient country. Even with a selfish objective, i.e. minimizing fatalities in the donor country, we find that the optimal policy is for the donor country to share vaccines across a broad range of realistic vaccination rates. This effect is intensified as epidemic coupling between nations increases. These results suggest that selfish objectives can lead to increased vaccine sharing. Furthermore, by donating vaccines the outbreak and deaths in a recipient nation can be reduced significantly.

The optimal policy of vaccine allocation is dependent on the epidemic coupling constant and the daily vaccination rates. We find that in the regime of high vaccination rates and low-to-moderate values of the epidemic coupling coefficient, the optimal sharing fraction is 0, supporting the results shown in [10]. However, as the vaccination rate decreases, the optimal policy is to share vaccines. We interpret this finding as follows: when vaccination rates are low and stockpiles are high, then a donor country is not effectively using its vaccine. Instead, a large part of its population could become infected before they can get vaccinated. Therefore it is better for a donor country to share vaccines with a recipient country. As a result, the recipient country begins to vaccinate individuals thereby reducing the cross-national force of infection due to coupling between the two countries. We find that vaccine sharing can increase to high levels in the limit of low per-capita vaccine rates and moderate between-nation coupling. We note that vaccine sharing remains optimal even in the limit of very low coupling when vaccination rates are sufficiently low.

The finding that selfish objectives leads to vaccine sharing policies can be extended to alternative, near-optimal policies. We propose a hybrid policy that involves sharing vaccines (e.g., 1/3 of the vaccine stock) even when the optimal policy is not to share. As vaccination rates decrease, this hybrid policy has small to negligible impacts on fatalities in the donor country while leading to significant reductions in fatalities in the recipient country. In certain cases it may be reasonable to adopt such a hybrid policy by a donor nation, especially if the emergence of new strains of the virus are incorporated in the dynamic model of the pandemic.

The model used for simulating the epidemic and evaluating vaccine sharing policies comes with caveats. The effects of infections in one nation on another is modeled and simulated through the use of an epidemic coupling constant rather than through more complex travel policies (e.g., [14, 15]). In searching for optimal sharing policies, we assume that both nations have equal vaccination rates given available stockpiles and similar efficiency of isolating infected cases. In reality, wealthy nations likely can vaccinate at a higher rate and are likely to be more efficient in case tracing and isolation than poorer countries. Finally, we have assumed that immunity does not wane over the time-scale of the optimization (i.e., approximately 1 year). In reality, immunity wanes due to intrinsic changes and the emergence of new variants [16, 17, 18]. Incorporating these realistic scenarios in the model and analysis should be targets of future research.

In summary, the optimization framework developed in this paper demonstrates the value of sharing of vaccines based on the selfish objective of minimizing fatalities in a donor country. Despite the selfish objectives, we find broad regimes (based on the vaccination rates and epidemic coupling constants) where sharing vaccines is both optimal for the donor country and leads to significant reductions in fatalities for both the donor and recipient countries. It is hoped that extensions of this modeling and optimization framework encourage re-evaluations of vaccine-sharing policies towards increased global cooperation.

## Methods

The dynamic model of the disease that we analyze consists of a SEIRV framework for each one of the two considered countries, connected by a force of infection describing the crossover rate of infection between them. The population of country *i* (*i* ∈ {*A, B*}) is divided into the following groups: susceptible, exposed, infectious, recovered, and vaccinated. The respective sizes of these groups are denoted by *S*_*i*_, *E*_*i*_, *I*_*i*_, *R*_*i*_, and *V*_*i*_, and additionally, the total number of fatalities is denoted by *D*_*i*_. The population vector of country *i* is denoted by *P*_*i*_ := [*S*_*i*_, *E*_*i*_, *I*_*i*_, *R*_*i*_, *V*_*i*_]^*⊤*^. Let *c*_*S*_, *c*_*E*_, *c*_*I*_, *c*_*R*_ and *c*_*V*_ denote the respective contact rates of the subpopulations *S*_*i*_, *E*_*i*_, *I*_*i*_, *R*_*i*_ and *V*_*i*_ in country *i*, where it is assumed for the sake of simplicity of presentation that each one of these quantities is identical for both countries. Define *c* as the vector of contact rates, *c* = [*c*_*S*_, *c*_*E*_, *c*_*I*_, *c*_*R*_, *c*_*V*_]^*⊤*^. Furthermore, let *η*_*I*_ be the measure of disease transmission effectiveness from an infectious person to an exposed individual [19], and let *κ* be the epidemic coupling constant capturing the reduced contact rate between the populations in countries A and B.

The dynamics of the two nations are coupled through their *force of infection* defined, in country *i*, as

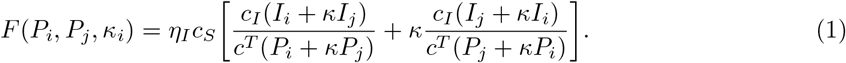

The term 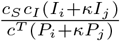 in the Right-Hand Side (RHS) of Eq. (1) is the probability that a susceptible individual from nation *i* comes in contact with an infected individual in country *i* regardless of the latter’s nation; note the role of the force of infection *κ* in attenuating *I*_*j*_. Similarly, the term 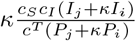 in the RHS of Eq. (1) represents the probability that a susceptible individual from nation *i* comes in contact with an infected individual at country *j* regardless of the national identity of the latter.

This notion of the force of infection extends the standard notion associated with a single country [20]. To see this, suppose that all the contact rates are equal to a baseline contact rate, *c*_*B*_, and *κ* = 0, namely the pandemics in the two nations are decoupled. Assuming relatively few fatalities per capita, the total population is approximately constant *N*, hence the force of infection can be approximated by *F* ≈ *η*_*I*_*c*_*B*_(*I/N*); this gives the standard SEIR model by defining *β = ηc*_*B*_ as the *baseline infection rate*.

Returning to the case discussed in this paper, assume that a person can be vaccinated only if in the susceptible state, and denote by *λ*_*i*_ the daily vaccination rate in country *i*. We assume that *λ*_*i*_ is dependent on vaccine availability in the following way. Given a constant *λ >* 0 representing the operational vaccination rate in either country, *λ*_*i*_ is defined as

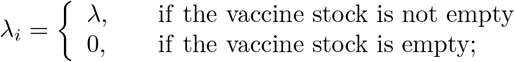

note that *λ* is assumed to be independent of the specific country where the vaccination is performed. Let the total vaccine-stock available before sharing be *V*_0_. At the start of the simulation, country A donates a fraction of its vaccine stock to B, denoted by *µ ∈* [0, 1]. Let 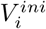 denote the initial vaccine stock in country *i*, thus 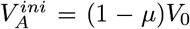 and 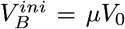. Furthermore, let *T*_*inf*_ be the infectious period; *T*_*inc*_ be the disease incubation period; and *ϕ* be the case fatality ratio for infected individuals. In the present case, the model for country *i* is given by the following system of differential equations,

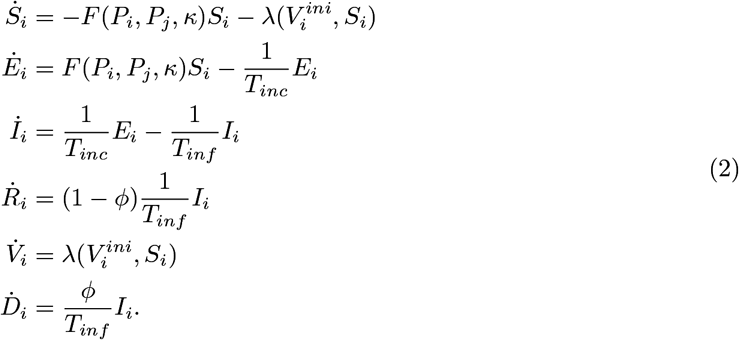

We assume that the contact rate for all the subpopulations except the infected group is equal to the baseline contact rate, *c*_*B*_, while for the infected subpopulation it is set to *c*_*B*_*/*2 to reflect the fact that infected people can be partially isolated.

The vaccine sharing event determines the respective available vaccine stocks in the two countries as functions of time throughout the simulation horizon. We formulate an optimization problem over all possible values of *µ ∈* [0, 1] with the aim of minimizing the fatalities in country A at the final time, *t*_*f*_. Defining the fatalities in country A at time t as *D*_*A*_(*t*), and the cost function as *J*(*µ*) = *D*_*A*_(*t*_*f*_), the optimization problem is

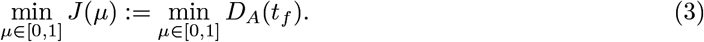

We solve this problem by a dynamic gradient-descent algorithm based on optimal control whose code, written in MATLAB, is available in https://github.com/WeitzGroup/vaccine allocation.

## Data Availability

All data produced are available online at https://github.com/WeitzGroup/vaccine_allocation

## Acknowledgments

We thank Benjamin Lopman, Eugenio Valdano, and Weitz Group team members for comments and feedback. This work was supported by grants from the Army Research Office (W911NF1910384) and National Institutes of Health (1R01AI46592-01).

### Appendix

**Table S1:** Parameters and initial conditions used in state equations (2). In the table above, *i* ∈ {*A, B*} and the initial conditions for both nations are the same.

| Parameter | Meaning | Value |
| --- | --- | --- |
| $\eta_I$ | Measure of disease transmission effectiveness | 0.1 |
| $T_{inc}$ | Mean incubation period | 4 days |
| $T_{inf}$ | Mean infectious period | 6 days |
| $\phi$ | Case fatality ratio of infected cases | 0.01 |
| $c_B$ | Baseline potentially infectious contact rate | 5/day |
| $S_i(0)$ | Initial susceptible population in country $i$ | $10^7 - 500$ |
| $E_i(0)$ | Initial exposed population in country $i$ | 0 |
| $I_i(0)$ | Initial infected population in country $i$ | 500 |
| $R_i(0)$ | Initial recovered population in country $i$ | 0 |
| $V_i(0)$ | Initial vaccinated population in country $i$ | 0 |
| $D_i(0)$ | Initial fatalities in country $i$ | 0 |
| $V_0$ | Total vaccines available to country A | $7 \times 10^6$ |
| $t_f$ | Time horizon | 360 days |

**Figure S1:**
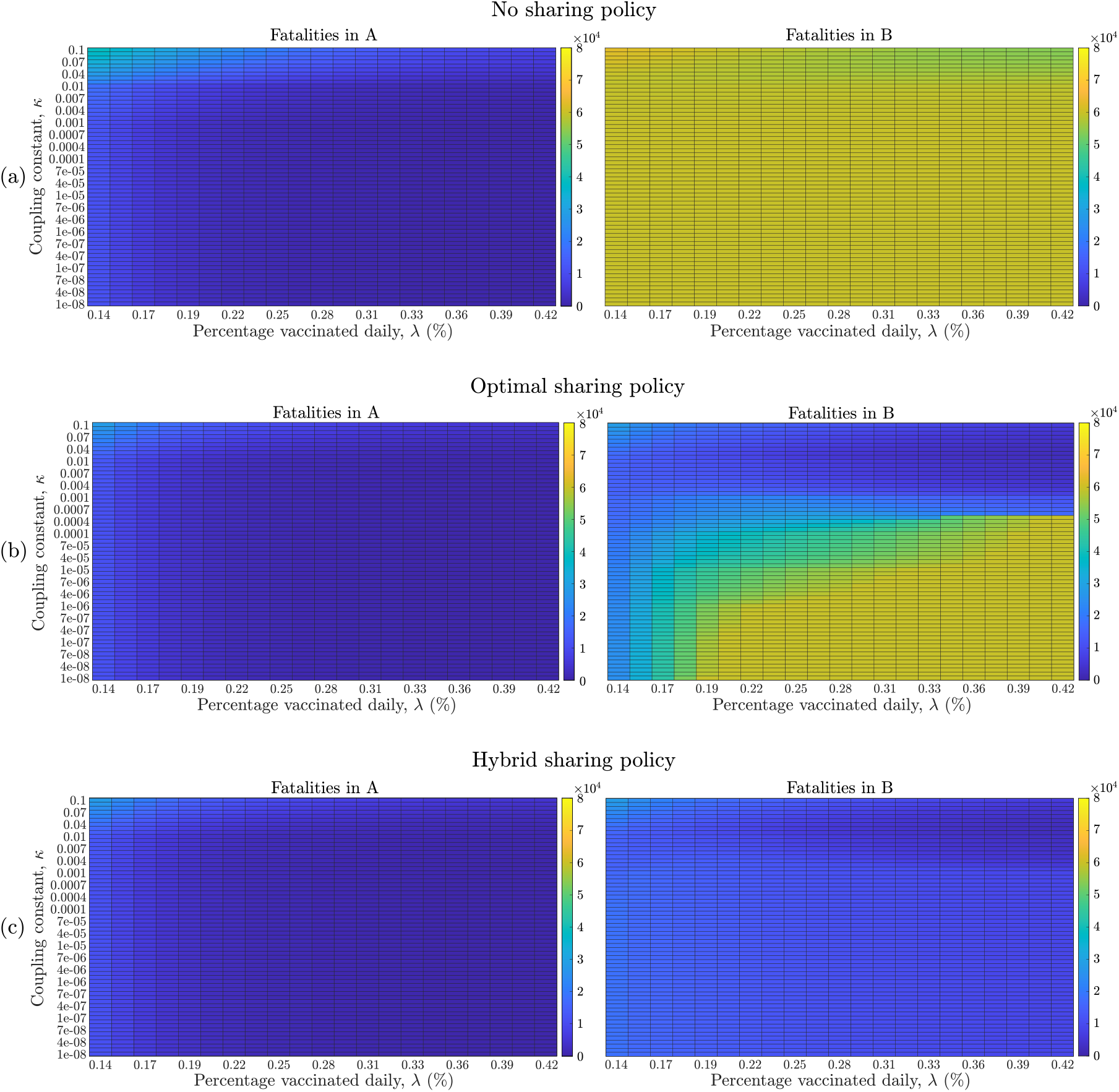
Fatalities in countries A and B when the **(a)** no-sharing policy, **(b)** optimal policy and **(c)** hybrid policy is implemented, over different coupling constants, *κ* ∈ [10^−8^, 10^−1^] and vaccination rates, *λ* (from 0.14% to 0.42% of the population daily). The optimal policy has the objective of minimizing fatalities in country A and the optimal sharing fraction is *µ*\*. The hybrid policy is a near optimal policy with a sharing fraction of 1/3 when *µ*\* ≤ 1*/*3 and equal to the optimal sharing fraction otherwise. The optimal policy shows significant reduction in fatalities in B when vaccination rate is low and coupling constant is high, when compared to the no-sharing policy. The hybrid policy results in major reduction in fatalities in B for all vaccination rates and coupling constants, when compared to the no-sharing policy. There is a slight increase in fatalities in A for the hybrid policy when the coupling constant is very small and the vaccination rate is high.

**Figure S2:**
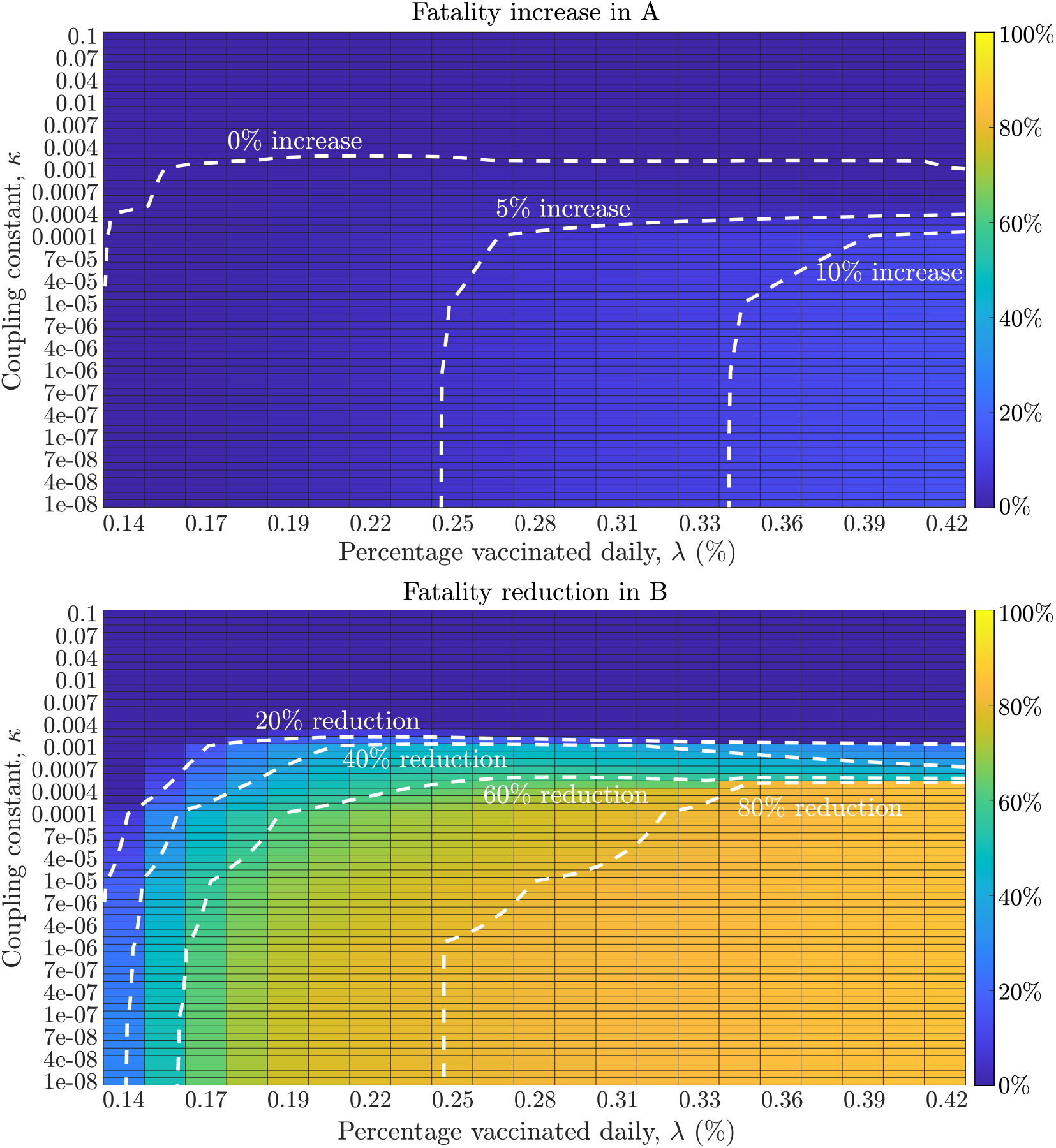
Percentage change in fatalities in countries A and B when comparing the hybrid policy with the optimal policy, over different coupling constants, *κ* ∈ [10^−8^, 10^−1^] and vaccination rates, *λ* (from 0.14% to 0.42% of the population vaccinated daily). The hybrid policy is a near optimal policy with a sharing fraction of 1/3 when *µ*\* ≤ 1*/*3 and equal to the optimal sharing fraction otherwise. For country A, an increase in fatalities in the range of [0%, 14%] is observed and contours are made for 0%, 5% and 10% increase in fatalities. For country B, a decrease of fatalities in the range of [0%, 85%] can be seen, with contours plotted for 20%, 40%, 60% and 80% fatality reduction. By relaxing the strict condition of minimizing fatalities in A, the near optimal solution provided by the hybrid policy shows significant fatality reduction in B when compared to the optimal policy. However, this comes at the cost of a small increase in fatalities in A in the regime of low coupling constant and high vaccination rate.

## Notes

### Competing Interest Statement

The authors have declared no competing interest.

### Funding Statement

This study was funded by grants from the Army Research Office (W911NF1910384) and National Institutes of Health (1R01AI46592-01)

